# Quarantine fatigue thins fat-tailed coronavirus impacts in U.S. cities by making epidemics inevitable

**DOI:** 10.1101/2021.01.07.21249366

**Authors:** Marc N. Conte, Matthew Gordon, Charles Sims

## Abstract

We use detailed location data to show that contacts between individuals in most U.S. cities and counties are fat tailed, suggesting that the fat tails documented in a small number of superspreading clusters are widespread. We integrate these results into a stochastic compartmental model to show that COVID-19 cases were also fat tailed for many U.S. cities for several weeks in the spring and summer. Due to epidemiological thresholds, fat-tailed cases would have been more prevalent if not for the gradual increase in contact rates throughout the summer that made outbreaks more certain.

---

Public officials have struggled to effectively control the spread of COVID-19. Fat tails in the distribution of global pandemic fatalities over the past two millennia (1) offer a possible explanation for these challenges. Fat-tailed damages across disease outbreaks limit the ability to learn and prepare for future outbreaks, as the central limit theorem slows down and fails to hold with infinite moments, so extreme events may be more likely than suggested by experience. We extend recent results showing fat tails in superspreading events (2, 3) to demonstrate the emergence and persistence of fat tails in contact distributions across the U.S. We then demonstrate an interaction between these contact distributions and community-specific disease dynamics that results in fat-tailed distributions of COVID-19 impacts (proxied by weekly cumulative cases) during the exact time when attempts at suppression were most intense.

## Results

We follow the “place-based” approach developed in (4), rather than proxying for contacts with time spent at home, and utilize the Safegraph weekly patterns dataset of visits to 6 million ‘Points-of-Interest’ (POIs) to estimate weekly contact distributions in 2,293 core-based statistical areas and rural counties (CBSAs) in the U.S. from January 27, 2020 to September 16, 2020 (SI Appendix and Dataset S1). Weekly estimates of the mean, variance, and shape parameter (*ξ*) of the Generalized Pareto distribution (GPD) all have predictive power for COVID-19 cases or deaths, demonstrating the relevance of our contacts measure (SI Appendix). Virtually all CBSAs (99%) have at least one week of a fat-tailed contact distribution with finite variance (0 < *ξ < 0*.*5*), and 35% of CBSAs have a fat-tailed distribution with infinite variance (*ξ > 0*.*5*) for at least 1 week.

Average contacts changed significantly during the epidemic due to defensive behavior and policy (5, 6). Our measure of contacts includes both POI and in-home contacts, which have been shown to be an important source of infectious-disease transmission (7). Generally, mean contacts fell, and the fraction of individuals staying completely at home increased significantly, but shape parameters spiked between week 10 (starting March 2) and week 13 (ending March 29) (Fig. 1A-B). For 25% of the CBSAs in our sample, the contact distribution entered infinite variance for at least one week as mean contacts were falling, suggesting that reductions in POI visits were concentrated in low-contact POIs, with some of these visits redirected to high-contact POIs in the tail. The histogram of contacts at NYC POIs (Fig. 1C) demonstrates a substantial reduction in visits to low-contact POIs between week 10 and week 13. However, the upper tail is thicker in week 13. This result holds across the U.S. and when breaking out different types of POIs, including restaurants, groceries, day care centers, and gyms (Fig. 1D). Restricted business hours and openings may have crowded more people into fewer locations, even as average POI visits fell. Following week 13, quarantine fatigue emerges in most CBSAs, leading to higher mean contacts and lower shape parameters.

**Fig. 1.**
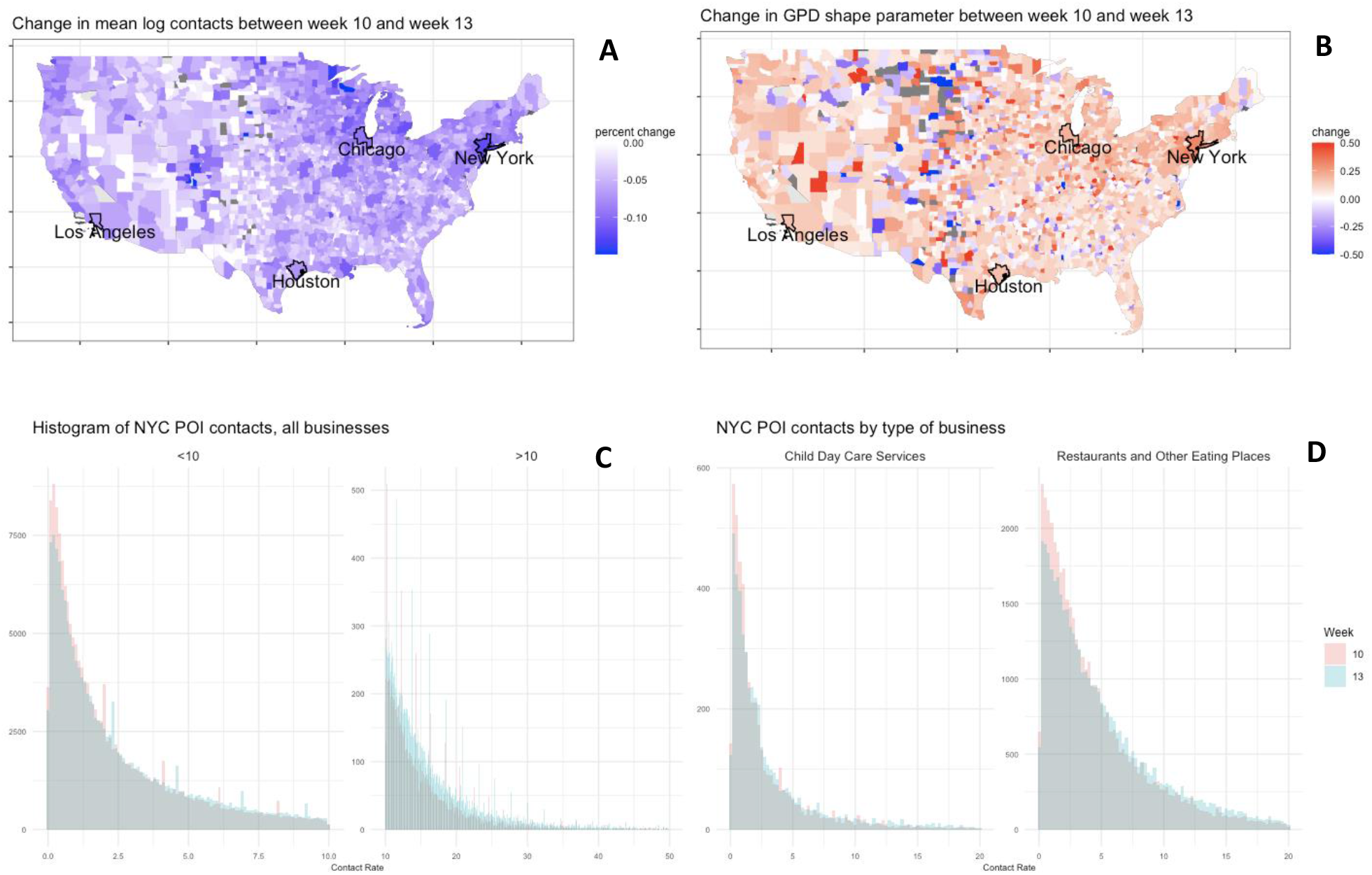
Mean and tail thickness of POI contact distributions. Mean contacts decrease during March (A), while estimated shape parameters increase in many places (B). In New York City, there are large reductions for POIs with low contacts, but an increase in POIs with high contacts during this period (C; tail split for ease of visualization). This pattern holds for restaurants and day care centers (D), among other types of businesses not pictured.

Our evidence of a general pattern of fat-tailed contact distributions across the U.S. suggests that fat tails in U.S. cases observed early in the outbreak (8) are due to city- and county-specific contact networks and epidemiological dynamics. To illustrate, we applied a two-step process to the four most populous U.S. cities: New York City, Los Angeles, Chicago, and Houston.

First, we fit an SIR model without demography to cases in each city: 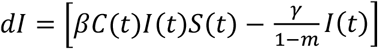 where *I*(*t*) and *S*(*t*) are the proportion of the city population *N* that is infected and susceptible respectively, *γ* is the city’s recovery rate and *m* is the city-specific probability an infected individual dies before recovering. Assuming frequency-dependent transmission, the per capita infectiveness is *βC*(*t*), capturing both the natural infectiveness of COVID-19, *β*, and the rate of contact between individuals in a city, *C*(*t*) (6). The contact rate follows a stochastic volatility model fit to our place-based contact distributions for each city: *dC* = *αCdt* + σ(*t*)*Cdz*_1_ where 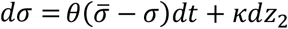 and *dz*_1_and *dz*_2_ are two independent Wiener processes (Fig. 2A) (SI Appendix). Our parameterized stochastic volatility model displays the quarantine fatigue observed in our contact distributions, *α* > 0, and implies contact rates are stochastic and fat tailed (SI Appendix). Most existing compartmental models account for contact rate stochasticity via Brownian motion processes that imply the contact rate distribution is thin tailed (e.g., 9). Ito’s lemma ensures the effective reproductive number, ℛ_0_(*t*) = *βC*(*t*)(1 − *m*)/*γ*, is also a fat-tailed random variable consistent with (2). Our stochastic volatility contact rate process leads to multiple waves of infected individuals *I(t)* with unpredictable timing and magnitude (Fig. 2B).

**Fig. 2.**
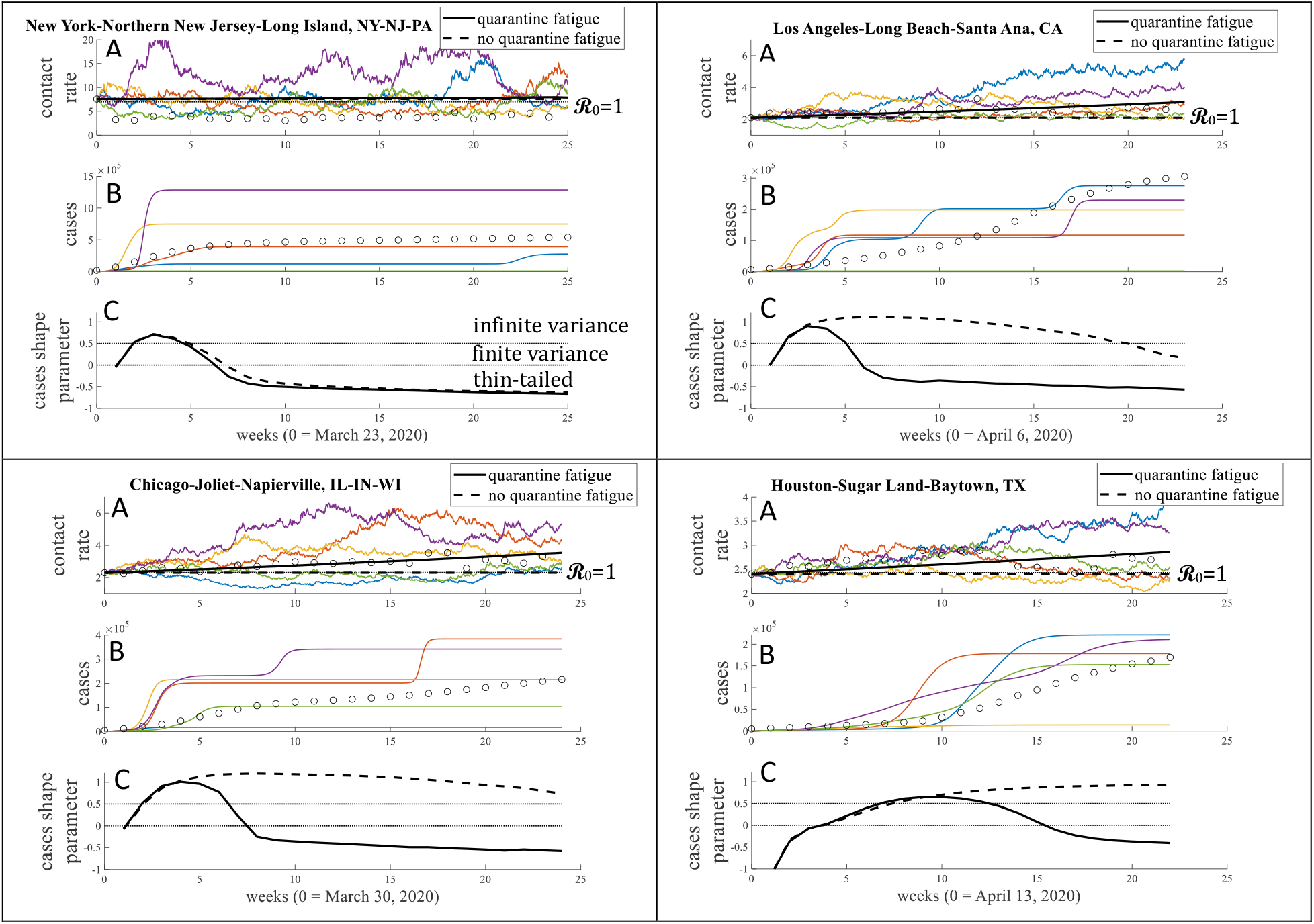
Duration of fat-tailed COVID-19 impacts. (*A*) Contact rates estimated from POI visits (circles) parameterize a stochastic process for contact rates (colored lines show 5 simulations of *dC* with quarantine fatigue) with expected increase over time due to quarantine fatigue (*E*[*dC*] > 0, solid black lines). (*B*) An SIR model fit to weekly cumulative cases (circles) is simulated using the stochastic contact rate process, yielding a time-varying distribution of cumulative cases (colored lines show 5 simulations of *dI* with quarantine fatigue). (*C*) GPD is fit to weekly distribution of cumulative cases with (*α* > 0; solid black lines) and without (*α* = 0; dashed black lines) quarantine fatigue.

Second, we perform 100,000 simulations of the parameterized stochastic SIR model and fit a GPD to the 100,000 simulated cumulative cases in each week, yielding weekly estimates of tail thickness of the COVID-19 impact distribution (Fig. 2C). New York City, Los Angeles, and Chicago all had fat-tailed cumulative case distributions in April and May. Houston, which experienced a delayed rise in cases, experienced fat-tailed impacts in late May, June, and July. All cities experienced at least 3 weeks with infinite variance impact distributions.

Public health policies and avoidance behaviors developed based on experiences during these months could be viewed as an overreaction if these impacts were mistakenly perceived as thin tailed, possibly contributing to reduced compliance, regulation, and the quarantine fatigue documented in our POI distributions. However, quarantine fatigue reduces the prevalence of fat tails by making an outbreak more certain. The gradual rise in contact rates due to quarantine fatigue means that more of the stochastic draws from the SIR model result in outbreaks. Greater certainty about an outbreak causes the shape parameter in cumulative cases to fall even as the mean in cumulative cases rises. Without quarantine fatigue (*α* = 0), ℛ_0_ hovers near 1 and more stochastic draws from the SIR model result in the epidemic being suppressed. Those stochastic draws resulting in an outbreak, which fell near the center of the distribution under quarantine fatigue, now appear more extreme, leading to a prolonged period of fat-tailed impacts (Fig. 2C). Thus, suppression of the disease can create fat tails due to a threshold in stochastic epidemiological dynamics (10).

## Discussion

In extreme value theory, fat tails confound efforts to prepare for future extreme events like natural disasters (e.g., 11) and violent conflicts (12) because experience does not provide reliable information about future tail draws. However, extreme events play out over time based on policy and behavioral responses to the event, which are themselves dynamically informed by past experiences. By unpacking the dynamics of an extreme event, we illustrate a tradeoff – fat tails in the distribution of COVID-19 impacts can be alleviated by quarantine fatigue. Quarantine fatigue thins the tails over time by normalizing case counts that seemed extreme early in the epidemic. If the case distribution is incorrectly perceived as thin tailed, public health policies and avoidance behaviors could be viewed as an overreaction, perpetuating quarantine fatigue and sharpening this tradeoff.

Policy-makers might avoid this tradeoff with targeted interventions at superspreader POIs in the tail of the contact distribution that could make the case distribution thin tailed, potentially avoiding the increase in mean cases created by quarantine fatigue. While fat-tailed contacts are associated with superspreaders (13), increased transmission (2, 3), and higher case numbers (14), they also signal targeted interventions may be highly effective if policy makers have access to the necessary information and a mandate to act decisively. Our place-based estimates of contact distributions aid in these efforts by showing the dynamics of individual movement as the outbreak progresses, which is quite costly to achieve in network models, forcing the assumption of static contact networks in many models (e.g., 15).

## Data Availability

We have a spreadsheet of CBSA-level contact distribution summary statistics available upon request of the authors. The link to the case data is below.

https://github.com/CSSEGISandData/COVID-19

## Notes

### Competing Interest Statement

The authors have declared no competing interest.

### Funding Statement

The authors have no external funding sources to report.

### Author Declarations

No IRB review required.

### Summary of Updates

author affiliations updated

