## Supplemental Information for "Quarantine fatigue thins fat-tailed coronavirus impacts in U.S. cities by making epidemics inevitable"

[1] Department of Economics, Fordham University, Bronx, NY 10458, ORCID: 0000-0003-2859-9863
[2] School of the Environment, Yale University, New Haven, CT 06511, ORCID: 0000-0001-8241-3754
[3] Howard H. Baker Jr. Center for Public Policy, University of Tennessee, Knoxville, TN 37996 ORCID: 0000-0002-0458-8249

*Place-based measures of contact distribution*

To generate our contact distributions, we rely on cell phone data from the Safegraph weekly patterns dataset, which aggregates data from 45 million mobile devices in the United States, and visits to 6 million 'Points-of-Interest' (POIs). We build contact distributions based on visits to each POI in the dataset as follows:

Contact = ADV*CR*DF*s

ADV is the average daily visitors to the point of interest during the week. CR is the contact radius, a proxy for the crowdedness of a POI, defined as some radius within which transmission could occur, expressed as a fraction of the POI's square footage. In practice we use 10 and 20 feet and find similar results. DF is the median dwell time in a given POI expressed as a fraction of the hours a POI is open – defined as the number of unique hours that at least 1 visitor was observed in the POI, and s is a scaling factor that scales up to the population size given the number of devices observed by Safegraph. This effectively makes our contact equivalent to the expected number of people that would come within a certain radius of a visitor to a given POI during the week in question, assuming visitors are equally likely to go anywhere within the POI at any time that it is open. Thus, the uniform mixing assumption of compartmental models holds within POIs in our model, but our approach allows for more complex contact patterns across POIs within a CBSA.

To generate a complete 'place-based contact distribution', we also need to account for contacts within the home, which have been shown to be an important source of transmission (1, 2). Using the Safegraph social distancing dataset, we calculate the average fraction of devices that remain completely at home each day during the week. We then make that fraction of the POIs in our distribution 'homes' and assign them a contact of the average household size in the CBSA.
We then calculate the sample mean and variance of these distributions, and estimate the shape parameter of the tail of the distribution using maximum likelihood on the upper 50$^{th}$ percentile of the POI distribution (3). We confirm these results using the mean-excess plots to estimate the shape parameter (4). We also do not estimate shape parameters for CBSAs with very few POIs (less than 21). We do this for 34 weeks for nearly 2,300 CBSAs resulting in parameters for more than 75,000 distributions.

*Correlations between contact distribution parameters and epidemiological dynamics*

We run regressions using the following specification:

$$Y_{i,t+1} = \beta_1 Y_{i,t} + \beta_2 \Omega_{i,t} + \beta_3 Y_{i,t} * \Omega_{i,t} + \beta_4 X_{it} + \mu_i + \gamma_t$$

$Y$ is the log of the outcome of interest, either Covid-19 cases or deaths per 100,000 population, in CBSA *i* during week *t*. For Covid-19 deaths and case data, we use the Johns Hopkins Covid 19 data repository from https://github.com/CSSEGISandData/COVID-19 (5). Given the large number of zeroes, we use the inverse hyperbolic sine transformation. $\Omega$ is a vector of variables describing the distribution of contacts, including the mean and variance of the log of POI contacts, and maximum likelihood estimated shape parameters. We also include these contact variables interacted with the lagged case or death rate, which tells us how their marginal effects change as cases/deaths increase. $\mu_i$ and $\gamma_t$ are CBSA and week fixed effects, and $X_{it}$ is a vector of control variables, including the fraction of devices observed in a CBSA relative to the months before the epidemic. Regressions are also weighted by CBSA population.

All regressions have high goodness-of-fit as measured by R squared, especially the regressions on case rates, and the coefficients on our contact distribution variables are highly significant, showing that changes in the mean, variance, and tail behavior of our constructed contacts all have predictive power.

These results are robust to changing the size of the lead on the outcome variable, alternative definitions of the contact radius, as well as breaking out stay at home behavior and household size separately, and constructing a contact distribution based solely on visits to POIs outside the home.

*SIR model with stochastic, fat-tailed contact rates*

To determine if the impacts of the outbreak are fat-tailed, we use a novel stochastic variant of the standard SIR framework, which tracks the numbers of susceptible, infected, and recovered individuals over the course of an infectious disease outbreak:

$$dS(t) = -\beta C(t)I(t)S(t)dt \qquad (1)$$

$$dI(t) = \left[\beta C(t)I(t)S(t) - \frac{\gamma}{1-m}I(t)\right]dt \qquad (2)$$

$$dR(t) = \gamma I(t)dt \qquad (3)$$

where $I(t)$, $S(t)$, and $R(t)$ are the proportion of the total population $N$ that is infected, susceptible, and recovered respectively and $\frac{\gamma m}{1-m}I(t)$ is the daily change in the percent of the population that dies due to COVID-19. Assuming frequency dependent transmission, the per capita infectiveness of COVID-19 is $\beta C(t)$ which captures both the natural infectiveness of the disease, $\beta$, and the rate of contact between individuals in a population $C(t)$ (6).

The contact rate is a stochastic variable reflecting the unpredictability of contacts between individuals captured in our place-based contact distributions. While human behavior makes contact rates inherently unpredictable (7-9), our place-based contact rate measures also highlight how a CBSA's mix of businesses and building stock also enable or discourage variability in contact rates. For example, a CBSA with more high-contact POIs, such as full-service restaurants, fitness centers, and cafes (10), will also exhibit greater variability in contacts between individuals. To convert the place-based measure of contact rate to a contact rate compatible with our compartmental model, we assume the contact rate in CBSA $i$,$C_i(t)$, is the expected contacts per person in CBSA $i$: $C_i(t) = \frac{1}{2}\left[\frac{\mu_{i,t}^2+v_{i,t}}{\mu_{i,t}} - 1\right]$ where $\mu_{i,t}$ and $v_{i,t}$ are the mean and variance of the place-based contact rate distribution respectively.

We use the resulting time-series of contact rates to estimate a stochastic volatility model for contact rates for the four most populous U.S. cities. Similar stochastic volatility models are used in the finance literature to model fat-tailed stock price distributions (11). To inform the specification of our model, we first performed an augmented Dickey-Fuller unit root test to determine whether the time series of contact rates is a random walk or trend stationary. For all 4 CBSAs, we fail to reject the null hypothesis of a unit root thus ruling out a variety of mean-reverting processes. Based on this finding:

$$dC_i = \alpha_i C_i dt + \sigma_i(t) C_i dz_{1i} \tag{4}$$

where $\alpha_i$ is a drift term, $\sigma_i(t)$ is a diffusion or variance term, and $dz_{i1}$ is the increment of a standard Wiener process which are independent across CBSAs. $\alpha_i < 0$ indicates that, on average, the contact rate is falling over time in CBSA *i* due to self-protection behaviors and policy (6), while $\alpha_i > 0$ indicates that the contact rate is generally rising over time in CBSA *I* due to quarantine fatigue (12). More unpredictable human behavior would manifest as a more variable contact rate and a larger value for $\sigma_i(t)$. If $\sigma_i(t)$ were a fixed parameter, contact rates would be log-normally distributed leading to thick-tailed contact rates, $\xi = 0$. However, all 4 CBSAs exhibited fat tails ($\xi > 0$) throughout the 34 weeks of our study. To allow for fat tails, the diffusion or volatility parameter is also stochastic and follows an arithmetic Ornstein-Uhlenbeck (or AR(1)) process:

$$d\sigma_i = \theta_i(\bar{\sigma}_i - \sigma_i)dt + \kappa_i dz_{i2} \tag{5}$$

where $\bar{\sigma}_i$ is the long-run average level of percent volatility in contact rates, $\theta_i$ is the speed of mean reversion, $\kappa_i$is the volatility in the percent volatility, and $dz_{i2}$ is a second independent Wiener process. The stochastic volatility model in equations 4 and 5 ensures $C_i(t)$ will be fat-tailed with finite variance and can exhibit infinite variance with certain parameter combinations (11). This is in contrast to many other models that account for stochasticity in contact rates by assuming $C_i$ is a normally distributed random variable (7, 8).

We estimate the drift term and the long-run average level of percent volatility as:

$$\widehat{\alpha_\iota} = \frac{1}{N}\sum_{t=1}^{N} log\left(\frac{C_{ti}}{C_{t-1i}}\right) + \frac{\widehat{\sigma_i}^2}{2}$$

$$\widehat{\sigma_i} = \sqrt{\frac{1}{N-1}\sum_{t=1}^{N}\left(log\left(\frac{C_{ti}}{C_{t-1i}}\right) - \frac{1}{N}\sum_{t=1}^{N} log\left(\frac{C_{ti}}{C_{t-1i}}\right)\right)^2}$$

Our estimates for the drift term range from 0.019 in Chicago to 0.002 in New York City indicating quarantine fatigue was most pronounced in Chicago and least pronounced in New York City. Our finding of quarantine fatigue is consistent with contact behavior in previous epidemics (12). Our estimates for the long-run average level of percent volatility range from 0.21 in New York City to 0.04 in Houston. These parameter values provide an indication of the degree of unpredictability in the contact in each city. For example, contact rates in New York City increased 0.2% each week with a 21% volatility around this trend.

Our weekly contact rate data do not provide enough observations to confidently estimate an AR(1) model needed to recover estimates of $\theta_i$ and $\kappa_i$. However, the relationship between these parameters and results from the augmented Dickey-Fuller test give us several clues about their relative magnitude. The unconditional standard deviation of volatility in our model is given by $\kappa_i/(2\theta_i)^2$. Thus, small values of $\kappa_i$ can have a large effect on the data generating process if the speed of mean reversion is low. Conversely, large values of $\kappa_i$ can have a small effect on the data generating process if the speed of mean reversion is fast. In each CBSA, we set $\kappa_i = \frac{1}{2}\bar{\sigma}_i$. The results of our augmented Dickey-Fuller test suggest $\theta_i$is relatively large to ensure the random walk properties of the data are preserved. Given $\kappa_i = \frac{1}{2}\bar{\sigma}_i$, we select values for $\theta_i$ in each CBSA that ensures consistent rejection of the null hypothesis of a unit root. These values imply that the half-life of a volatility shock ranges from 2 weeks ($\theta_i$=19.22) in New York City to 6 weeks ($\theta_i$=3.82) in Houston. Our parameter values imply that the unconditional standard deviation of volatility ranges from 0.02 in New York City (8% of its mean) to 0.008 in Houston (18% of its mean). Our results do not qualitatively change under various combinations of $\theta_i$ and $\kappa_i$ values that results in rejection of the unit root null hypothesis.

Due to the population sizes in our four cities, we assume contact rate stochasticity (a form of environmental stochasticity) dominates demographic stochasticity. SIR models with demographic stochasticity and constant contact rates produce approximately normal distributions for the number of infected individuals when $\mathcal{R}_0 > 1$ and the total population, $N$, is sufficiently large (13, 14). However, the distribution of infected individuals will not be normal when $\mathcal{R}_0$ is close to 1 (13, 14). This result is qualitatively similar to ours in that the distribution of cumulative cases is fat-tailed when $\mathcal{R}_0$ is near 1 and becomes thin tailed as $\mathcal{R}_0$ moves further from 1. However, we describe a different mechanism for this phenomenon rooted in quarantine fatigue that shifts the distribution of contact rates over time. Given sufficient data, our stochastic contact rate SIR model can accommodate demographic stochasticity using a diffusion approximation to a continuous time Markov chain process similar to (9).

We use weekly case counts for each of the 4 CBSAs to estimate the epidemiological parameters $\beta_i$, $\gamma_i$, and $m_i$ that minimize the sum of squared errors between the weekly observed case counts in a CBSA and the expected path of $I_i(t)$ for that CBSA over a period of 34 weeks.

We then perform 100,000 simulations of the fitted stochastic SIR model in Matlab using the Euler approach to approximate continuous-time stochastic processes. To alleviate concerns about under reporting of cases early in the outbreak, we set the initial condition for the simulation equal to the week where the percent infected in a CBSA first exceeds 0.0005. This initial condition is as early as March 23, 2020 (New York City) and as late as April 13, 2020 (Houston). We then calculate the cumulative sum of cases and deaths for each simulation as a proxies for the damages incurred by COVID-19. In each week, we fit a GPD to the 100,000 simulated cumulative cases and deaths yielding a weekly estimate of the thickness of the tails (shape parameter, $\xi_i$) for the cumulative impacts of COVID-19 in each CBSA. We use maximum likelihood to estimate the two parameters of the GPD.

The stochastic epidemiological system in equations 1-5 assumes the current proportion of the population that is infected, susceptible, and recovered is known but that the future course of the outbreak is unknown due to the inability to predict future behaviors that determine the contact rate. This approach ensures that the fat tails we find are due to extreme draws in individual behavior due to factors such as superspreader locations. While uncertainty in current cases and deaths (i.e., state uncertainty) are important sources of uncertainty to consider when developing testing protocols, and other public health responses, they can lead to fat tails in cumulative cases due to intermittent changes in testing efforts and individual willingness to volunteer for testing. For example, an "extreme draw" for COVID-19 cases would be expected prior to Thanksgiving and Christmas as individuals seek out testing as part of quarantine procedures prior to visiting family. The uncertainty in our distributions does not reflect this type of state uncertainty.
